# Sonographic Diaphragm Abnormalities are an Unexpectedly Frequent Feature of Long COVID Outpatients with Unexplained Dyspnea and Fatigue

**DOI:** 10.1101/2022.06.29.22277054

**Authors:** Prabhav P. Deo, Joseph I. Bailey, Alexandra S. Jensen, Ellen Farr, Meghan Fahey, Matthew Isherwood, Keerthana Chakka, Lisa F. Wolfe, Ishan Roy, Marc A. Sala, Colin K. Franz

## Abstract

**Introduction/Aims:** The primary aim of this study is to define the sonographic diaphragm phenotype of Long COVID rehabilitation outpatients with non-specific dyspnea and fatigue. We analyzed patients referred from a pulmonary post-COVID clinic that were lacking a specific cardiopulmonary diagnosis for their symptoms. Additionally, we report the functional outcomes of subset of patients who completed an outpatient cardiopulmonary physical therapy program.

**Methods:** This was a retrospective cohort study (n = 58) of consecutive patients referred for neuromuscular ultrasound assessment of diaphragm muscle using B-mode technique. Patients were recruited from a single academic hospital between February 25, 2021 and November 22, 2022.

**Results:** Sonographic abnormalities were identified in 57% (33/58) of patients, and in the vast majority of cases (33/33) was defined by a low diaphragm muscle thickness. Thinner diaphragm muscles are correlated with lower serum creatinine and creatine kinase values, but there was no association with markers of systemic inflammation. Thirty three patients participated in outpatient cardiopulmonary physical therapy that included respiratory muscle training, and 75.8% (25/33) had documented improvement.

**Discussion:** In the outpatient rehabilitation setting, patients with Long COVID display low diaphragm muscle thickness, but intact muscle contractility, with surprising frequency on neuromuscular ultrasound. We speculate this represents a form of disuse atrophy. Also, these patients appear to have a favorable response to cardiopulmonary physical therapy that includes respiratory muscle training.

## Introduction

Chronic functional impairments reported in survivors of the most severe coronavirus disease 2019 (COVID-19) who require inpatient rehabilitation to recover frequently have a neuromuscular underpinning.^1,2^ The extent of neuromuscular contribution to persistent symptoms observed after less severe COVID-19 is not well understood. According to the Centers for Disease Control and Prevention, Long COVID, also known as “post-COVID conditions” or “post-acute sequelae of SARS CoV-2 infection”, is defined as a wide range of new, returning, or ongoing health problems four or more weeks after initial infection.^3^ Persistent symptoms are present in more than half of COVID-19 survivors 6 months after recovery,^5^ with a frequency ranging from 10-35% in those with mild disease.^6^ Fatigue, dyspnea, and cough are among the most commonly reported symptoms in long COVID with a prevalence of 31%-63%, 18%-47%, and 12%-25%, respectively.^7^

After severe COVID-19 infection, Farr et al. used neuromuscular ultrasound to demonstrate impaired diaphragmatic contractility as a contributor to persistent dyspnea and fatigue in the setting of inpatient rehabilitation.^2^ However, the sonographic findings in patients with Long COVID in the outpatient rehabilitation setting has not yet been reported. We hypothesized that in this less severely affected population, diaphragm muscle structural abnormalities contribute to their persistent respiratory symptoms. Here we performed a retrospective analysis on Long COVID patients referred for neuromuscular rehabilitation assessments including diaphragm ultrasound after no clear cardiopulmonary etiology was identified. Additionally, we compared these results to outcomes from a cardiopulmonary physical therapy program.

## Methods

This is a retrospective analysis of the first 58 consecutive Long COVID patients with suspected neuromuscular respiratory weakness referred to a single center (Shirley Ryan AbilityLab, Chicago, IL) for neuromuscular ultrasound and consultation by a physiatrist who is board certified in physical medicine and rehabilitation, and neuromuscular medicine, as well as holds the neuromuscular ultrasound certificate of added qualification (American Board of Electrodiagnostic Medicine). All patients developed persistent dyspnea and/or fatigue after PCR-confirmation (47/58) or clinical diagnosis of COVID-19. No patients in this cohort required inpatient rehabilitation services after their acute infection, most (42/58) were never hospitalized, and none of them required invasive ventilation for their acute illness. The patients were evaluated between February 25, 2021 and November 22, 2022. Prior to rehabilitation referral, patients were evaluated by a pulmonologist within a comprehensive COVID care model at an academic affiliate (Northwestern Memorial Hospital, Chicago, IL). In addition to history and physical examination, CT scans were obtained in 39 patients (67%), pulmonary function tests were obtained in 49 patients (84%), and 43 patients (74%) underwent 6-minute walk testing. If this testing did not reveal a specific pulmonary parenchymal or pulmonary vascular etiology that fully explained the degree of post-infectious dyspnea observed, the patient was referred to outpatient rehabilitation clinic for diaphragm muscle ultrasound.

All patients underwent point of care ultrasound and 53 patients (91%) underwent electrodiagnostic testing that included phrenic nerve conduction studies plus or minus a limited needle electromyography exam of the biceps brachii and/or vastus lateralis muscle to screen for myopathic patterns. Previously published healthy control normative data set (n=150)^8^, as well as our prior data set of patients with more severe COVID infection who were evaluated during inpatient rehabilitation stay (n = 21)^2^ were used as a comparative populations. The diaphragm muscle was assessed on a portable ultrasound system (Xario 200, Canon Medical Systems USA Inc., Tustin, CA) with either a 5.0 to 18.0-MHz linear array (18L7) or 1.8-to 6.0-MHz convex array (6C1) selected to optimize image clarity on the basis of individual characteristics including body habitus in B-mode as previously described.^2^ Each hemi-diaphragm was identified in the zone of apposition, and thickness was measured at functional reserve capacity (FRC) and total lung capacity (TLC). Normative values have been established for diaphragm muscle thickness at FRC (≥ 0.14 cm) and thickening ratio (TLC/FRC muscle thickness; > 1.2) at the 8^th^ and 9^th^ intercostal space.^8^ Approval was obtained from Northwestern University IRB (STU23625789).

Cardiopulmonary physical therapy intervention was offered to all patients, which consisted of cardiovascular conditioning, with continuous monitoring of SpO2, heart rate and rate of perceived exertion. Patients were prescribed a respiratory muscle trainer, allowing for resistance during inspiration and expiration to complete as a home exercise program five times daily. In addition, patients were educated on shortness of breath, diaphragmatic breathing strategies and habit forming strategies to continue with cardiovascular conditioning and breathing program independently post-discharge.

Since this data did not have a normal distribution as determined by the Shapiro–Wilk test, the Wilcoxon Signed Rank test (nonparametric, dependent values) was utilized to compare mean left and right hemi-diaphragm measurements. The Mann-Whitney U test (nonparametric, independent values) was used to compare less severe COVID rehabilitation clinic outpatients (current data) with Farr et al. data set from more severe COVID rehabilitation unit inpatients.^2^ Pearson correlation coefficient (r) was employed to correlate sonographic findings with laboratory data and functional outcomes. Ultrasound data is displayed throughout the results section as mean ± standard deviation.

## Results

The baseline characteristics of the 58 patients assessed in outpatient rehabilitation medicine clinic after COVID-19 infection, including demographic data, hospital course, spirometry, and laboratory testing are reported in Table 1. The sonographic diaphragm thickness measured at FRC and thickening ratios of the left and right hemi-diaphragms are displayed in Figure 1A, C. The mean left hemi-diaphragm thickness was 0.17 ± 0.06 cm compared to 0.18 ± 0.06 cm on the right (p=0.86). The mean thickening ratio was 2.1 ± 0.5 for the left hemi-diaphragm compared to 2.1 ± 0.6 for the right hemi-diaphragm (p=0.91). Fifty seven percent (33/58) of patients had at least one sonographic value below the lower limit of normal, and in the vast majority of abnormal cases (32/33) it was reduced hemi-diaphragm muscle thickness at FRC (< 0.15 cm). Sonographic findings in COVID rehabilitation outpatients compared to more severe COVID rehabilitation inpatients can be seen in Figure 1B, D. There was a statistically trend towards a smaller left and right hemi-diaphragm thickness in the outpatient rehabilitation population compared to our prior inpatient rehabilitation data set^2^ (left 0.20 ± 0.08 cm, p=0.05; right 0.21 ± 0.08 cm, p=0.06). The left and right hemi-diaphragm thickening ratio was greater in the outpatient population compared to the inpatient population (left 1.3 ± 0.3, p<10^e-5^; right 1.3 ± 0.3, p<10^e-5^).

**Table 1.** Baseline characteristics and laboratory values of Long COVID outpatient rehabilitation patients. Results are reported as Median value (range of values). Abbreviations: Intensive care unit (ICU); Forced expiratory volume (FEV1); Forced vital capacity (FVC); Diffusing capacity for carbon monoxide (DLCO); Total Lung Capacity (TLC); C-reactive protein (CRP); Creatine kinase (CK); Absolute neutrophil count (ANC); Absolute leukocyte count (ALC); 6 minute walk test (6MWT).

| Baseline characteristics and laboratory values of Long COVID outpatient rehabilitation patients |  |  |
| --- | --- | --- |
| Age (n=58) | 46 (24 - 78) |  |
| Sex M:F (n=58) | 14:44 |  |
| BMI (n=53) | 29.4 (14.56– 63.9) |  |
| Comorbidities (n=58) | Hypertension | 13.8% (8/58) |
|  | Diabetes mellitus and pre-diabetes mellitus | 12.1% (7/58) |
|  | Hyperlipidemia | 3.4% (2/58) |
|  | Asthma and/or COPD | 17.2% (10/58) |
|  | Cancer | 3.4% (3/58) |
|  | Acquired immunodeficiency | 5.2% (3/58) |
|  | Stroke | 1.7% (1/58) |
|  | OSA | 8.6% (5/58) |
|  | Allergies/Allergic Rhinitis | 5.2% (3/58) |
|  | Depression | 10.3% (6/58) |
|  | Anxiety | 13.8% (8/58) |
| Hospitalized (n=58) | 27.6% (16/58) |  |
| ICU (n=58) | 3.4% (2/58) |  |
| Days from COVID diagnosis to ultrasound scan (n=58) | 359 (65- 765) |  |
| FEV1% (n=49) | 90 (34 - 122) |  |
| FVC% (n=49) | 92 (44 - 130) |  |
| FEV/FVC (n=49) | 79 (58 - 94) |  |
| DLCO% (n=49) | 88 (50 - 115) |  |
| TLC% (n=48) | 97 (60 - 132) |  |
| CRP mg/L (n=40) | 4.1 (0.6 – 18) |  |
| CK units/L (n=32) | 89 (30 – 190) |  |
| D-Dimer ng/mL (n=41) | <150 | 51.4% (24/41) |
|  | 268 (153 – 911) | 29.7% (17/41) |
| Albumin g/L (n=33) | 4.4 (2.6 – 4.8) |  |
| Bicarbonate mEq/L (n=54) | 25.8 (18 – 31) |  |
| ANC/ALC (n=33) | 2.6 (0.6 – 7.7) |  |
| 6MWT m (n=43) | 427 (94 - 738) |  |

**Figure 1.**
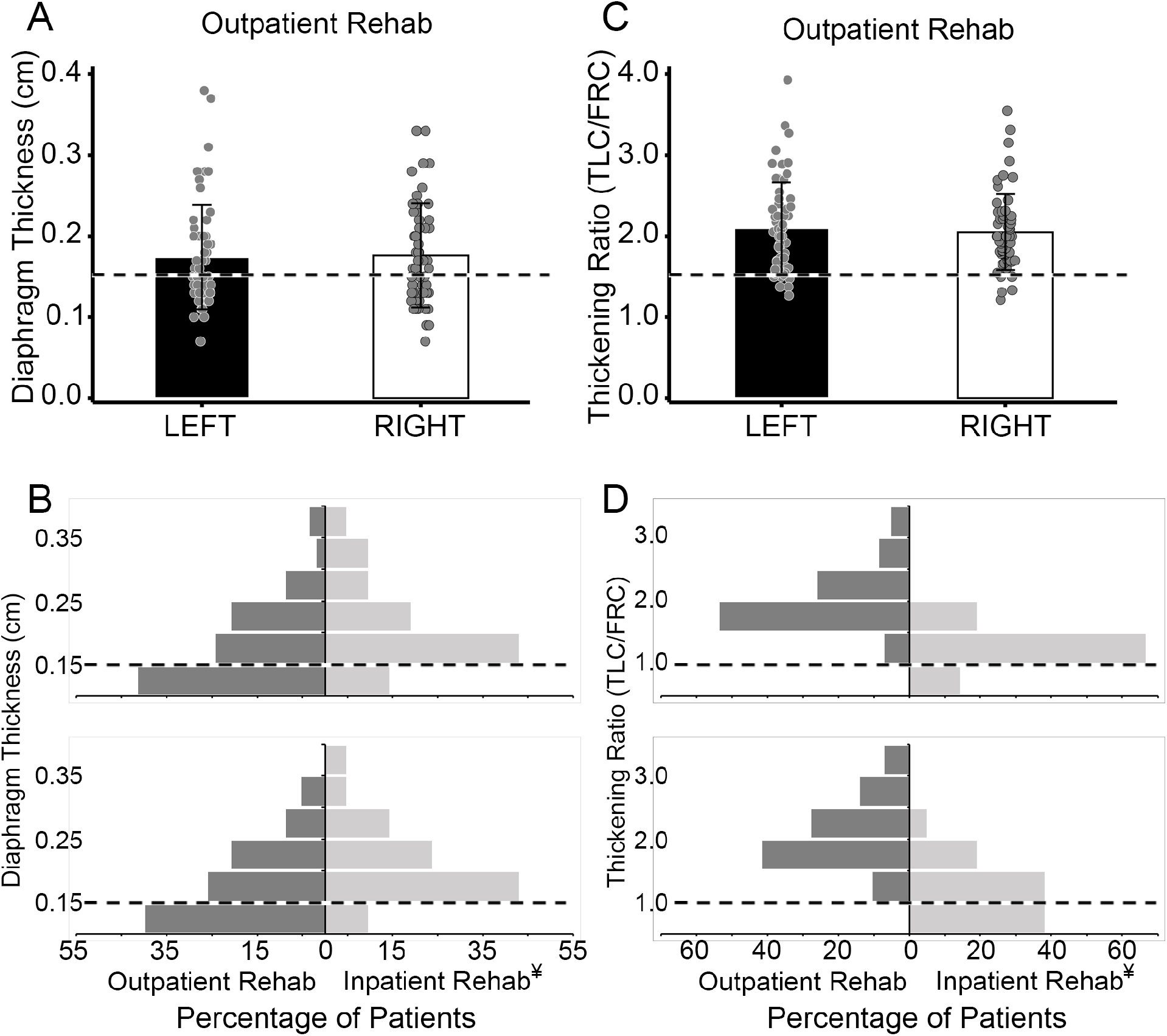
Diaphragm sonographic measurements in Long COVID rehabilitation outpatients including comparison to prior cohort of COVID rehabilitation inpatients. (A) The diaphragm muscle thickness, assessed at functional residual capacity (FRC), is plotted for the left (black bar) and right (white bar) sides of each patient (n=58). (B) Histogram comparing left (top) and right (bottom) diaphragm muscle thickness (at FRC) for Long COVID rehabilitation outpatients (dark grey bars) versus our previously published COVID rehabilitation inpatients (light grey bars). (C) The diaphragm muscle thickening ratio, which is the ratio of the muscle thickness taken at total lung capacity (TLC) over FRC, is plotted for the left and right side of each patient. (D) Histogram comparing left (top) and right (bottom) diaphragm thickening ratio (TLC/FRC) for Long COVID rehabilitation outpatients (dark grey bars) versus our previously published COVID rehabilitation inpatients (light grey bars). The dotted horizontal lines indicate lower limit of normal for each parameter. ¥, inpatient COVID rehabilitation data reproduced from Farr et al. 2021.

### Electrodiagnostic Findings

Limited needle electromyography exam was performed to screen for myopathic changes of the biceps brachii and/or vastus lateralis muscles on at least one side in most patients (50/58). There were no electrodiagnostic findings to suggest an active or chronic myopathy noted (e.g. fibrillation potentials, positive sharp waves, small amplitude and/or polyphasic motor unit potentials). None of the patients were clinically diagnosed with phrenic neuropathies, but 6/58 had absent phrenic nerve conduction responses despite normal diaphragm thickening ratios that was clinically interpreted as a false positive result (10.3%), which is a known limitation of surface recordings of evoked diaphragm motor response.^9^

### Functional Outcomes

A subset of COVID-19 outpatients accepted a referral for outpatient cardiopulmonary physical therapy (n=33) with an emphasis on respiratory muscle training. Patients attended a mean of 5.8 ± 3.9 training sessions. For the patients who did not enroll in physical therapy (n=25), the main reason was preclusive distance from their residence to the outpatient therapy office in downtown Chicago (19/25). Some may have pursued outpatient cardiopulmonary rehabilitation closer to their residence at an outside facility, but no records of this were available to include. Of those who did enroll, 11 patients who had 6-minute walk test as an objective measurement before and after completion of physical therapy, with a median change of +36 meters, and 63.6% (7/11) of patients showing improvement (Figure 2A). In one patient who was noted to regress on 6-minute walk, this change was attributed to aggravation of pre-existing knee osteoarthritis felt to be unrelated to cardiopulmonary physical therapy. In terms of subjective changes noted by chart review of physical therapy notes, 75.8% (25/33) had a documented subjective benefit, 3.0% (1/33) subjectively worsened, and 6.1% (7/33) had no improvement or not tolerate therapy (Figure 2B). Reasons charted for inability to tolerate physical therapy include fatigue, chronic pain, or preexisting medical comorbidities such as arthritis.

**Figure 2.**
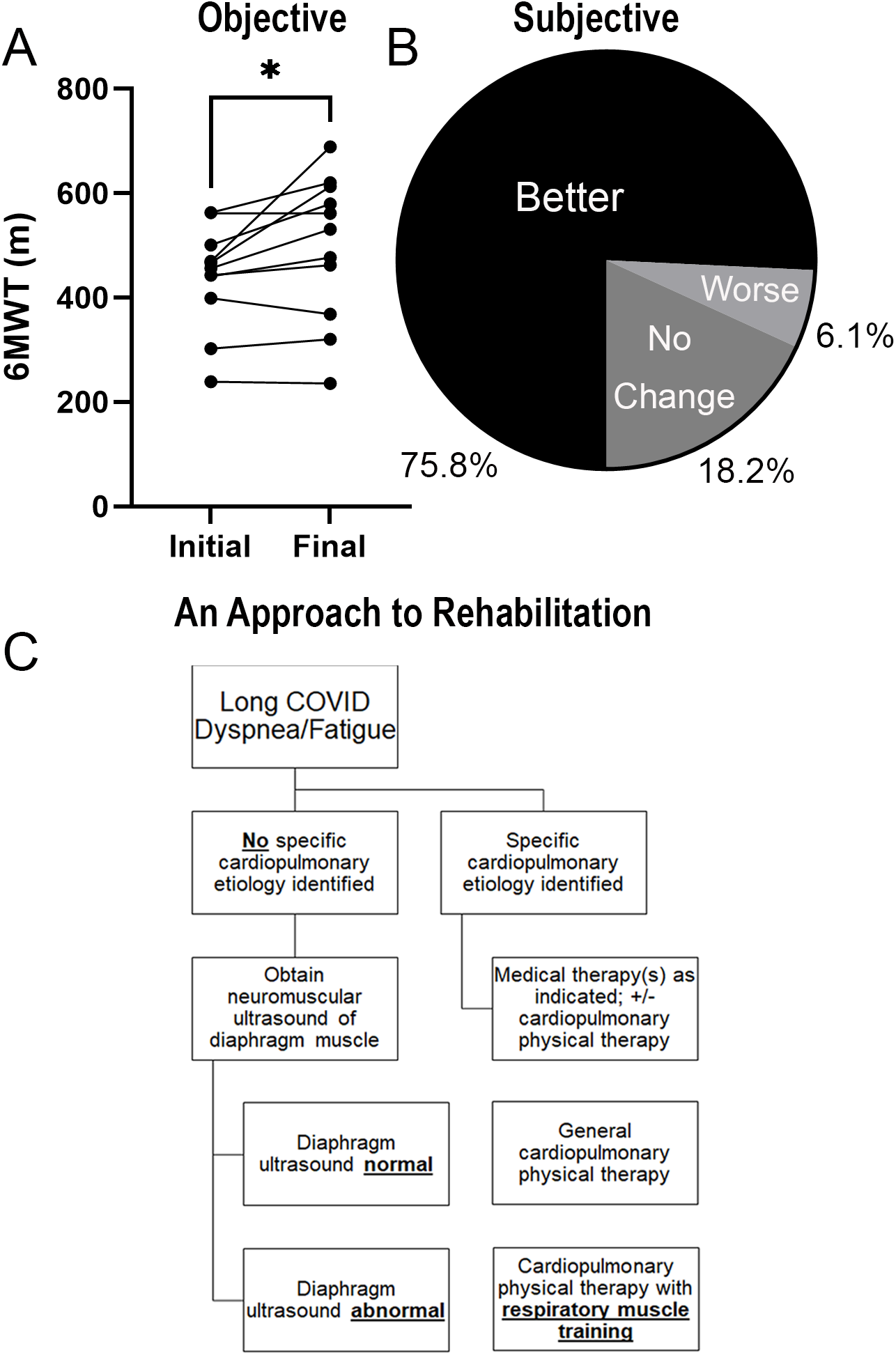
Functional outcomes for subset of Long COVID rehabilitation outpatients who participated in cardiopulmonary physical therapy program including our suggested approach for making cardiopulmonary rehabilitation referrals. (A) The objective improvement (n=10) as assessed by six minute walk test (6mwt) after completion of outpatient cardiopulmonary therapy (median = +36.0 meters; p=0.0244). (B) The percentages of Long COVID patients (n=33) who had documented (subjective) improvements after participating in outpatient cardiopulmonary therapy for their non-specific pattern of dyspnea and/or fatigue. (B) Cardiopulmonary physical therapy referrals may be considered for non-specific symptoms of dyspnea and/or fatigue in all Long COVID patients, but consider emphasis on respiratory muscle training exercises specifically for patients who have an objective abnormality on diaphragm muscle ultrasound

### Pulmonary Testing

Pulmonary function testing was obtained for patients at the clinical discretion of the providing physician. Spirometry, whole body plethysmography, nitrogen washout, and diffusing capacity of the lung for carbon monoxide (DLCO) were obtained. One patient was unable to tolerate lung volume assessment. Six minute walk testing was also performed in clinic prior to rehabilitation referral for 43 patients by trained medical assistants. Pulmonary function testing and 6 minute walk test results are presented in Table 1. Forced vital capacity, forced expiratory volume in 1 second, TLC, and DLCO are presented as % predicted. Briefly, the median FVC was 92% (range 44-130%). The median forced expiratory volume in 1 second (FEV1) was 90% (range 34-122%). The median FEV1/FVC ratio was 79% (range 58-94%). The median TLC was 97% (range 60-132%). The median diffusing capacity for carbon monoxide (DLCO) was 88% (range 50-115%).

CT scans were obtained for patients at the clinical discretion of the pulmonary medicine physician. Clinical radiology reports were reviewed by the research team. Of the 39 patients who underwent CT scans to evaluate their dyspnea, the most common abnormality noted were pulmonary nodules, which were identified in 11 (28.2%) of patients. One nodule was 8mm, but all others were <5mm in size. Five patients (12.8%) had small areas of ground glass opacities and 3 (7.7%) were found to have band-like opacities, concerning for areas of fibrosis. None of the patients had an elevated hemi-diaphragm (0%) by radiography.

### Laboratory Markers of Muscle Wasting Disease

To address the possibility that this cohort is impacted by inflammatory muscle wasting, we examined C-reactive protein (CRP), CK, D-Dimer, bicarbonate, albumin, neutrophil to lymphocyte ratio, creatinine, albumin, and prognostic nutritional index (Supplemental Figure 1). The relationship between these serum-based markers of muscle wasting disease and functional assessments (6 minute walk test, pulmonary function testing, or diaphragm thickness) was assessed through a Pearson correlation matrix. While no inflammatory based markers significantly correlated with any outcomes, BMI, serum creatinine and CK had significant, positive correlations with diaphragm thickness. BMI also had positive correlations with CRP and neutrophil to lymphocyte ratio.

## Discussion

This study defines a distinct Long COVID diaphragm phenotype in the outpatient rehabilitation setting for patients with persistent respiratory symptoms not accounted for by intrinsic cardiopulmonary pathology. In an unexpectedly high percentage of cases (55%) these Long COVID patients presented with decreased diaphragm muscle thickness (atrophy), but intact (57/58 patients) diaphragm muscle thickening ratio (contractility). This distinguishes them from our prior study on more severe COVID-19 survivors who were assessed while in the inpatient rehabilitation setting and predominantly had impaired diaphragm thickening ratio (67%) rather than reduced muscle thickness (18%) on sonographic exam.^2^ The mechanism for Long COVID diaphragm atrophy is unclear, but notably we did not find serological markers of persistent inflammatory or cachexia-like state correlated with atrophy (FRC thickness) of the diaphragm muscle to account for this effect (Supplemental Table 1). Encouragingly, 75% of the sub-group of Long COVID patients (n=33) that accepted referrals for outpatient cardiopulmonary rehabilitation had documented improvements on chart review (Figure 2A-B).

General neuromuscular causes of diaphragm dysfunction include motor neuron disease, advanced polyneuropathy, myopathy, generalized muscle wasting (e.g. cachexia), and phrenic mononeuropathy.^10,11^ In patients with severe COVID-19 infection, other proposed mechanisms include ventilator-induced diaphragm dysfunction and direct viral infiltration of SARS-CoV-2 through angiotensin converting enzyme 2 (ACE2).^12,13^ In our cohort of predominantly non-hospitalized patients (72%), there was no clinically diagnosed cases of phrenic mononeuropathy, polyneuropathy, or electrodiagnostic evidence of a myopathy. Recently a study of 467 patients hospitalized for COVID-19 pneumonia demonstrated that 3.2% (15/467) developed a new onset hemi-diaphragm elevation diagnosed by chest x-ray, however here we found no such cases (0/39) suggesting phrenic neuropathy in severe COVID-19 may be more a consequence of the critical illness itself rather than COVID-19 infection per se.^14^

Additionally, while the atrophy was likely not due to an active inflammatory state, the positive correlation between diaphragm thickness, serum creatinine, and CK implies generalized loss of muscle mass may contribute to this phenotype. This is extrapolated from the use of creatinine as a proxy marker of muscle mass in other neuromuscular disease states.^15,16^ Based on the activity intolerance reported by many of our Long COVID patients, muscle disuse atrophy should be considered as a potential explanation. This may explain why a clear majority of our patients responded favorably to cardiopulmonary physical therapy (75%). This rehabilitation framework was proposed by Severin and colleagues early in the pandemic,^18^ and in addition to the current results it is supported by a large meta-analysis showing positive effects of pulmonary rehabilitation on dyspnea, and quality of life.^19^ A small percentage of our cohort did not tolerate physical therapy, which raises the question of how best to approach treatment of their functional deficits and whether this status could change spontaneously over time allowing for successful intervention in the future.

Although other studies have used diaphragm ultrasound to characterize patients with severe COVID either acutely^2,20,21^ or in outpatient follow up,^22^ but little is known about diaphragm muscle structural changes in Long COVID especially in non-hospitalized patients that make up the majority of our current cohort (72%). Given the high rates of fatigue and dyspnea in people living with Long COVID^7^, this study contributes by identifying a structural abnormality (reduced diaphragm muscle thickness) that physical therapy can be specifically designed to directly address (Figure 2C). Limitations of our study include the relatively small sample size, as well as the retrospective and single center study design. In addition, quantitative functional outcome data pre- and post-COVID infection and physical therapy intervention was limited.

Future research on diaphragm dysfunction in Long COVID should consider prospective study design with repeated diaphragm function measurements. To better assess the full impact of cardiopulmonary physical therapy, a randomized controlled trial with intent to treat analysis would be desirable.

## Conclusion

In this retrospective cohort study, we were able to identify evidence of sonographic diaphragm abnormalities in more than half of the Long COVID patients referred to our outpatient rehabilitation clinic with non-specific dyspnea and fatigue (57%). In all but one case, the abnormality was sonographic muscle atrophy rather than grossly impaired diaphragm muscle contractility. The pathophysiologic mechanism of this finding is unclear, but we hypothesize it may be a form of disuse muscle atrophy due to the lack of association with systemic markers of inflammation. Ideally baseline diaphragm ultrasound scans would be available for patients, but given how difficult this would be to achieve prospectively, a future study with serial diaphragm ultrasounds representing states of acute infection and long term follow up would be informative. Our results suggest that patients with persistent respiratory symptoms after COVID-19 infection, which is not accounted for by an intrinsic cardiopulmonary etiology, should be considered for referrals for (i) neuromuscular ultrasound of diaphragm muscle, and (ii) outpatient rehabilitation medicine evaluation. If referred for cardiopulmonary physical therapy, it appears more likely than not, that patients who are able to participate will experience improvement in symptoms and function.^17^

## Data Availability

All data produced in the present study are available upon reasonable request to the authors.

## Acknowledgements

CKF acknowledges the generous support of the Belle Carnell Regenerative Neurorehabilitation fund.

---

**Supplemental Figure 1.**
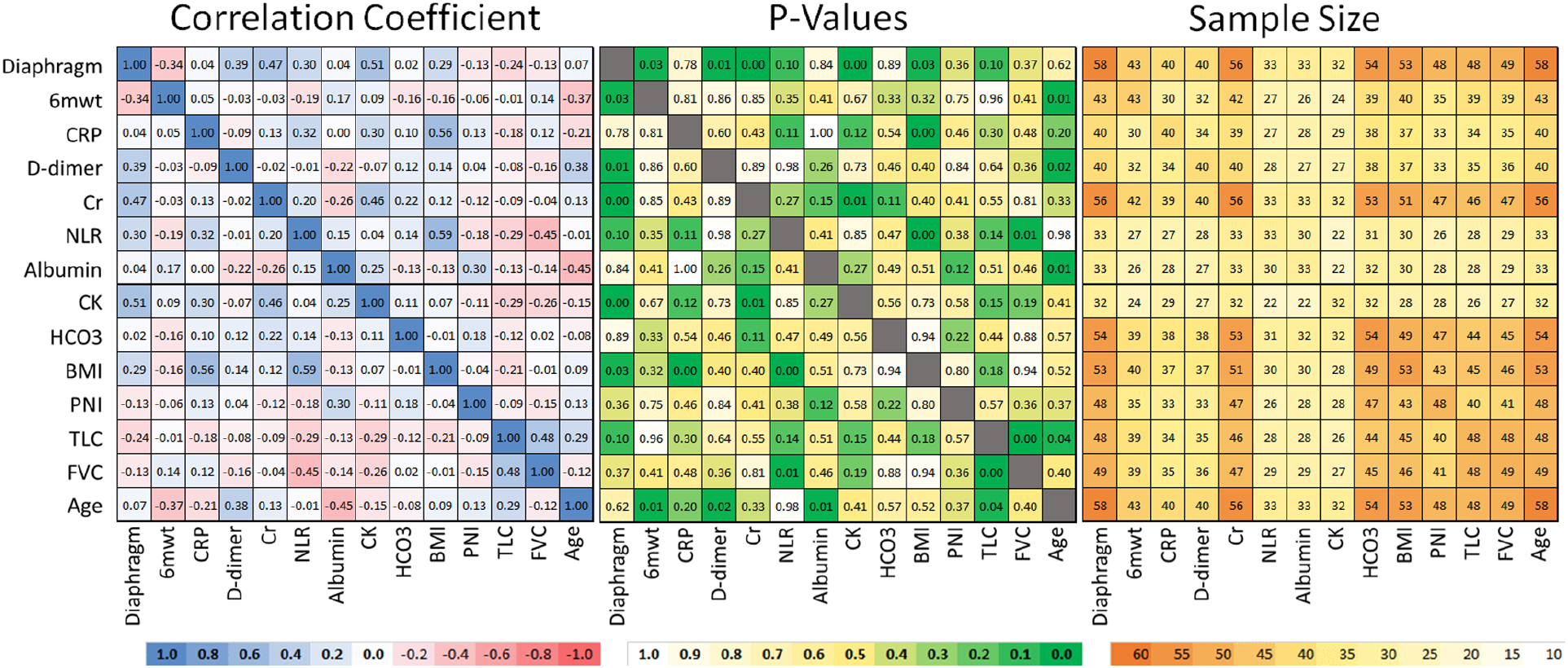
Heat map of Pearson correlation coefficient (left) and corresponding p-values (middle) and sample sizes (right). Selected parameters include diaphragm thickness at functional residual capacity (Diaphragm), distance walked on six minute walk test (6mwt), serum C-reactive protein (CRP), serum D-dimer, serum Creatinine (Cr), serum neutrophil lymphocyte ratio (NLR), serum albumin, serum creatine kinase (CK), serum bicarbonate (HCO3), body mass index (BMI), prognostic nutritional index (PNI), total lung capacity (TLC), forced vital capacity (FVC), and Age.

## References

1. Cabanes-Martinez L, Villadoniga M, Gonzalez-Rodriguez L, et al. Neuromuscular involvement in COVID-19 critically ill patients. Clin Neurophysiol. Dec 2020;131(12):2809–2816. doi:10.1016/j.clinph.2020.09.017

2. Farr E, Wolfe AR, Deshmukh S, et al. Diaphragm dysfunction in severe COVID-19 as determined by neuromuscular ultrasound. Ann Clin Transl Neurol. Aug 2021;8(8):1745–1749. doi:10.1002/acn3.51416

3. Evaluating and Caring for Patients with Post-COVID Conditions: Interim Guidance. Accessed 04/13/2022, 2022.

4. Ayoubkhani D, Bermingham C, Pouwels KB, et al. Trajectory of long covid symptoms after covid-19 vaccination: community based cohort study. BMJ. May 18 2022;377:e069676. doi:10.1136/bmj-2021-069676

5. Groff D, Sun A, Ssentongo AE, et al. Short-term and Long-term Rates of Postacute Sequelae of SARS-CoV-2 Infection: A Systematic Review. JAMA Netw Open. Oct 1 2021;4(10):e2128568. doi:10.1001/jamanetworkopen.2021.28568

6. van Kessel SAM, Olde Hartman TC, Lucassen P, van Jaarsveld CHM. Post-acute and long-COVID-19 symptoms in patients with mild diseases: a systematic review. Fam Pract. Jan 19 2022;39(1):159–167. doi:10.1093/fampra/cmab076

7. Aiyegbusi OL, Hughes SE, Turner G, et al. Symptoms, complications and management of long COVID: a review. J R Soc Med. Sep 2021;114(9):428–442. doi:10.1177/01410768211032850

8. Boon AJ, Harper CJ, Ghahfarokhi LS, Strommen JA, Watson JC, Sorenson EJ. Two-dimensional ultrasound imaging of the diaphragm: quantitative values in normal subjects. Muscle Nerve. Jun 2013;47(6):884–9. doi:10.1002/mus.23702

9. Boon AJ, Sekiguchi H, Harper CJ, et al. Sensitivity and specificity of diagnostic ultrasound in the diagnosis of phrenic neuropathy. Neurology. Sep 30 2014;83(14):1264–70. doi:10.1212/WNL.0000000000000841

10. McCool FD, Manzoor K, Minami T. Disorders of the Diaphragm. Clin Chest Med. Jun 2018;39(2):345–360. doi:10.1016/j.ccm.2018.01.012

11. Roberts BM, Ahn B, Smuder AJ, et al. Diaphragm and ventilatory dysfunction during cancer cachexia. FASEB J. Jul 2013;27(7):2600–10. doi:10.1096/fj.12-222844

12. Guarracino F, Vetrugno L, Forfori F, et al. Lung, Heart, Vascular, and Diaphragm Ultrasound Examination of COVID-19 Patients: A Comprehensive Approach. J Cardiothorac Vasc Anesth. Jun 2021;35(6):1866–1874. doi:10.1053/j.jvca.2020.06.013

13. Shi Z, de Vries HJ, Vlaar APJ, et al. Diaphragm Pathology in Critically Ill Patients With COVID-19 and Postmortem Findings From 3 Medical Centers. JAMA Intern Med. Jan 1 2021;181(1):122–124. doi:10.1001/jamainternmed.2020.6278

14. Law SM, Scott K, Alkarn A, Mahjoub A, Mallik AK, Roditi G, Choo-Kang B. COVID-19 associated phrenic nerve mononeuritis: a case series. Thorax Aug 2022;77(8):834–838. doi: 10.1136/thoraxjnl-2021-218257.

15. Chio A, Calvo A, Bovio G, et al. Amyotrophic lateral sclerosis outcome measures and the role of albumin and creatinine: a population-based study. JAMA Neurol. Sep 2014;71(9):1134–42. doi:10.1001/jamaneurol.2014.1129

16. Thongprayoon C, Cheungpasitporn W, Kashani K. Serum creatinine level, a surrogate of muscle mass, predicts mortality in critically ill patients. J Thorac Dis. May 2016;8(5):E305–11. doi:10.21037/jtd.2016.03.62

17. Belli S, Balbi B, Prince I, et al. Low physical functioning and impaired performance of activities of daily life in COVID-19 patients who survived hospitalisation. Eur Respir J. Oct 2020;56(4) doi:10.1183/13993003.02096-2020

18. Severin R, Arena R, Lavie CJ, Bond S, Phillips SA. Respiratory Muscle Performance Screening for Infectious Disease Management Following COVID-19: A Highly Pressurized Situation. Am J Med. Sep 2020;133(9):1025–1032. doi:10.1016/j.amjmed.2020.04.003

19. Chen H, Shi H, Liu X, Sun T, Wu J, Liu Z. Effect of Pulmonary Rehabilitation for Patients With Post-COVID-19: A Systematic Review and Meta-Analysis. Front Med (Lausanne). Feb 2022;9:837420. doi:10.3389/fmed.2022.83742020.

20. Boussuges A, Habert P, Chaumet G, Rouibah R, Delorme L, Menard A, Million M, Bartoli A, Guedj E, Gouitaa M, Zieleskiewicz L, Finance J, Coiffard B, Delliaux S, Brégeon F. Diaphragm dysfunction after severe COVID-19: An ultrasound study. Front Med (Lausanne). Aug 2022; 9:949281. doi: 10.3389/fmed.2022.949281.

21. Franz CK, Murthy NK, Malik GR, Kwak JW, D’Andrea D, Wolfe AR, Farr E, Stearns MA, Deshmukh S, Tavee JO, Sun F, Swong KN, Rydberg L, Cotton RJ, Wolfe LF, Walter JM, Coleman JM 3rd, Rogers JA. The distribution of acquired peripheral nerve injuries associated with severe COVID-19 implicate a mechanism of entrapment neuropathy: a multicenter case series and clinical feasibility study of a wearable, wireless pressure sensor. J Neuroeng Rehabil. 2022 Oct 8;19(1):108. doi: 10.1186/s12984-022-01089-1.

22. J, Friedrich J, Regmi B, Geppert J, Jörn B, Kersten A, Giannoni A, Boentert M, Marx G, Marx N, Daher A, Dreher M. Diaphragm dysfunction as a potential determinant of dyspnea on exertion in patients 1 year after COVID-19-related ARDS. Respir Res. Jul 2022; 23(1):187. doi: 10.1186/s12931-022-02100-y.

